# The Burden of Chikungunya and Onyong’nyong Viral Infections in Uganda: A Hospital-Based Sentinel Surveillance Study

**DOI:** 10.1101/2025.07.19.25331648

**Authors:** Doris Nyamwaya, Nyamai Mutono, John Kayiwa, Julius Lutwama, Augustine Masinde, SM Thumbi, Martin Antonio, George Warimwe, Pontiano Kaleebu

## Abstract

**Background:** Chikungunya and Onyong’nyong viruses are mosquito-borne alphaviruses endemic to Africa, causing febrile illness with severe joint pain. While Chikungunya virus is known for its potential to cause life-threatening complications in neonates and vulnerable populations, Onyong’nyong virus remains vastly underreported due to weak surveillance and diagnostic overlap. As a result, Onyong’nyong virus’s true burden and epidemic potential remain dangerously underestimated.

**Methods:** We conducted sentinel hospital-based surveillance in Uganda to assess the occurrence, geographical distribution, and clinical manifestation of Chikungunya and Onyong’nyong viruses. Serological and molecular diagnostic methods were employed to detect active and recent infections. Epidemiological data were analyzed to characterize clinical manifestations and determine risk factors for severe disease.

**Results:** A total of 2,756 serum samples were collected from febrile patients across 63 districts in Uganda between 2018 and 2022. While no Chikungunya infection was detected by PCR, two cases tested positive for Onyong’nyong virus. Serological evidence of recent alphavirus infection was found in 8.79% of samples, with peak prevalence in 2020. Most positive cases reported fever, joint and muscle pain, and other non-specific symptoms. PRNT analysis revealed stronger neutralizing antibody responses to Onyong’nyong virus than Chikungunya virus, suggesting ongoing but under-recognized Onyong’nyong virus transmission across Uganda.

**Conclusion:** This study underscores the endemic nature of Onyong’nyong virus in Uganda and the need for enhanced surveillance, improved diagnostic capacity, and targeted public health interventions. These findings will inform clinical management strategies and contribute to preparedness against future outbreaks.

## Introduction

Chikungunya virus (CHIKV) and Onyong’nyong virus (ONNV) are antigenically related mosquito-borne alphaviruses of the *Togaviridae* family that cause recurrent outbreaks of febrile illness. Both viruses were originally identified in Africa^1^, and though ONNV has not been detected outside the continent the geographic range of CHIKV is now nearly global, including South America, Asia and Europe ^2^.

CHIKV is primarily transmitted by *Aedes* mosquitoes and infection typically presents with a sudden onset of fever, and debilitating joint pain ^3^. Other symptoms include headache, rash, myalgia, nausea, and, in some cases, prolonged joint symptoms or neurological complications ^4–6^. ONNV is transmitted by *Anopheles* mosquitoes and shares overlapping symptoms with CHIKV infection but is characterized by low-grade fever, marked arthritis affecting large joints, post-cervical lymphadenitis, and a generalized rash ^7,8^. A distinguishing feature of ONNV is the occasional “saddleback” fever pattern, in which symptoms subside and then recur after several days; a presentation less commonly observed with CHIKV ^9,10^. Despite these clinical distinctions, both infections are frequently misdiagnosed due to their symptomatic similarity with malaria, dengue, and other endemic febrile illnesses, leading to underestimation of their true burden^11^.

CHIKV has gained global attention following its re-emergence in 2004 and subsequent outbreaks across Africa, Asia, and the Americas ^12–14^. In contrast, ONNV remains under-recognized despite its historical outbreak significance in East Africa ^8,15–17^. Two major epidemics have been documented: the first between 1959 and 1962, affecting more than two million people across nine African countries, and the second in southern Uganda in 1996, which resulted in attack rates as high as 68% ^7,16,18,19^. Although existing evidence suggests that ONNV may be more widespread in East Africa than currently appreciated, surveillance and research have disproportionately focused on CHIKV ^20–22^. In this study, we leveraged an existing hospital-based sentinel surveillance system to estimate the relative contributions of CHIKV and ONNV to the burden of febrile illness in Uganda.

## Methods

### Sampling

This study was conducted as part of the Uganda Virus Research Institute (UVRI) national arbovirus surveillance program under the Uganda Ministry of Health (MoH) routine health care delivery and surveillance system for early warning of diseases, which monitors causes of acute febrile illness (AFI) through hospital-based sentinel surveillance sites. We analyzed serum samples archived between January 2018 and December 2022. Samples were collected from patients presenting to health facilities in 63 districts across Uganda **Figures 1 & 2** and stored in UVRI biobank.

**Figure 1:**
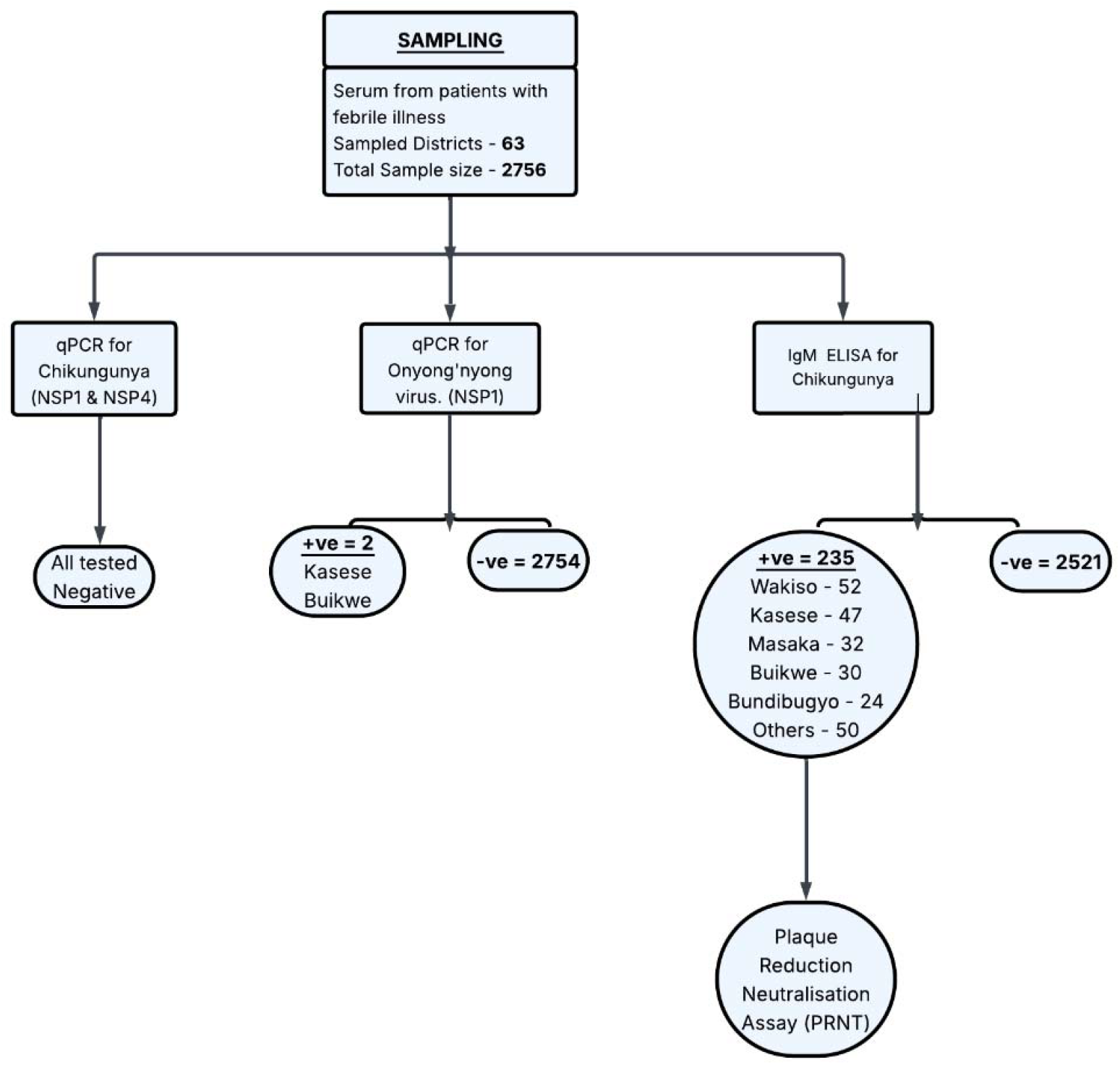
Consort diagram illustrating sampling and screening outcome for CHIKV and ONNV infections among febrile patients in Uganda.

**Figure 2:**
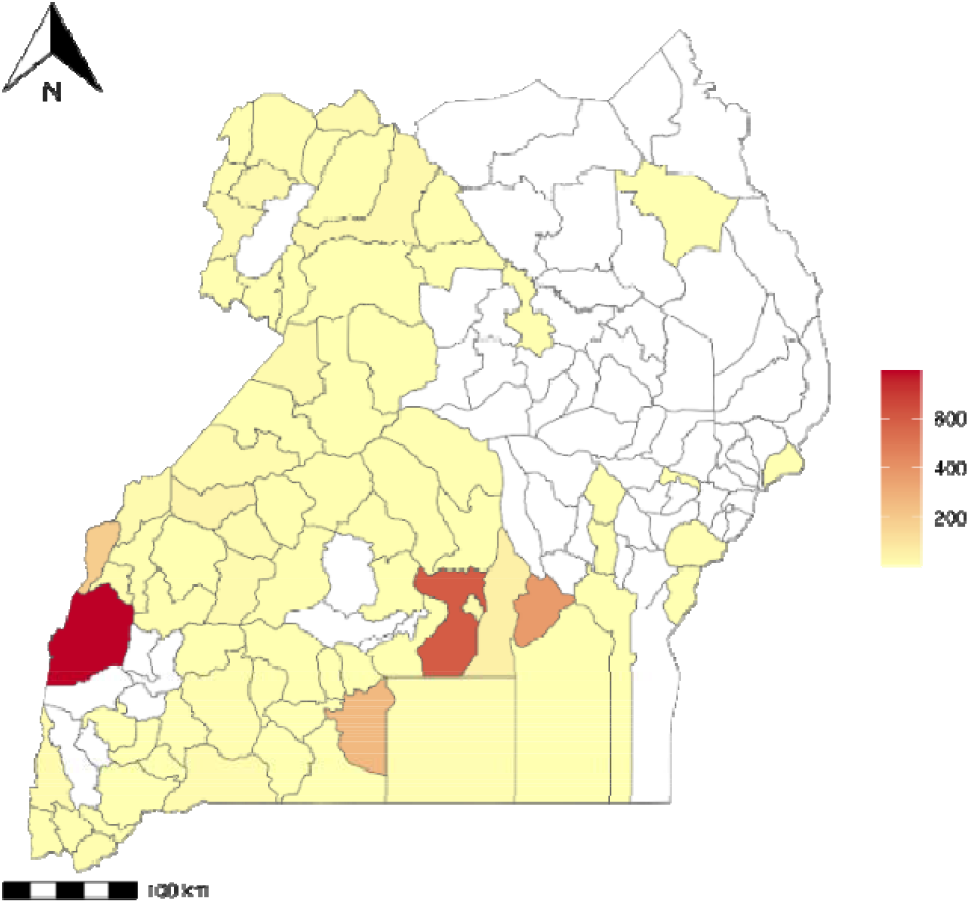
Geographic distribution of districts in Uganda where enrolled patients resided, based on hospital-based sentinel surveillance for chikungunya and Onyong’nyong virus infections (2018-2022)

### Participants and sample collection

Patients were eligible if they presented with an axillary temperature ≥37.5°C or reported a febrile illness lasting 2–7 days without a confirmed alternative diagnosis. Serum samples were collected, stored in liquid nitrogen at sentinel sites, and transported monthly to the UVRI Arbovirology Laboratory in Entebbe. At UVRI, samples were aliquoted for serological testing, virus isolation, and long-term storage. All available archived samples collected during the surveillance period were included in the analysis.

### Real time Reverse Transcription Polymerase Chain Reaction (RT-qPCR)

Total RNA was extracted from 140 μL of sampled serum using the QIAmp Viral RNA Mini Kit (Qiagen, Hilden, Germany), following the manufacturer’s instructions^23^. Detection of CHIKV or ONNV infection was based on the presence of viral RNA using RT-qPCR. For CHIKV, published primer-probe sets targeting the non-structural proteins 1 (nsP1) and 4 (nsP4) were used, while ONNV detection relied on a primer-probe set targeting the nsP1 region Error! Reference source not found.. The RT-qPCR reactions were carried out in 10 μL volumes using the Applied Biosystems™ TaqMan™ Fast Virus 1-Step Master Mix Kit, with final concentrations of 0.8 mM for primers, 0.2 mM for the probe, and 5 μL of RNA template. Amplification was performed on a 7500 Real-Time Fast PCR System (Applied Biosystems), with the following cycling conditions: reverse transcription at 50 °C for 5 minutes, initial denaturation at 95 °C for 20 seconds, followed by 45 cycles of 95 °C for 3 seconds and 60 °C for 30 seconds (data acquisition). A sample was considered positive if the cycle threshold (Ct) value was below 40. Positive controls included viral RNA extracted from cultured CHIKV R92082 and ONNV MP303 isolates, while negative controls consisted of RT-PCR master mix with PCR grade water as a template.

### Enzyme-Linked Immunosorbent Assay (ELISA)

Screening for acute and recent CHIKV infections in the study samples was carried out using the CHIKjj Detect IgM ELISA kit from InBios International (Seattle, Washington, USA) as per manufacturer’s instructions. This commercial kit is a sandwich ELISA designed to detect human IgM antibodies specific to the CHIKV E2/E1 envelope glycoproteins. However, due to potential cross-reactivity with other alphaviruses such as ONNV, positive ELISA results require confirmation through a virus neutralization test.

### Plaque Reduction Neutralization Test (PRNT)

This assay is based on the principle that neutralizing antibodies, which are produced as part of the immune response during infection, block virus attachment to host cells and inhibit infection. In the PRNT, a known amount of virus is incubated with test serum to assess the serum’s ability to neutralize the virus, with reference controls included for comparison. The endpoint is defined as the serum dilution at which 90% of viral plaques are reduced, indicating effective neutralization.

To perform the PRNT assay, serum samples were first heat-inactivated at 56°C for 30 minutes, then serially diluted two-fold in BA-1 diluent (10X M199 Hank’s Salts without L-Glutamine, 5% Bovine Serum Albumin, 1M TRIS-HCL pH 7.5, L-Glutamine, Sodium bicarbonate 7.5%, 100X penicillin/streptomycin, 1000X fungizone in sterile water). Each diluted serum (100 µL) was mixed with 100µL of BA-1 containing approximately 200 plaque-forming units (PFU) of the respective virus (CHIKV R92082 or ONNV MP303). The serum– virus mixtures were then added to confluent Vero cell monolayers in 12-well plates and incubated at 37°C (with 5% CO_2_) for 1 hour. The first overlay medium (comprising Miller’s 2X Teast Extract-Lactalbumin hydrolysate medium, 10X Earle’s Buffered Salts Solution, 2% fetal bovine serum, 1000X fungizone. 1000X gentamycin, and 2% low-melting agarose) was added, 3ml per well, and allowed to solidify for 30 minutes at room temperature. The plates were incubated at 37°C with 5% CO_2_ for 1 day. To stain the cell monolayers, neutral red dye mixed with second overlay (described as above) was added, 2ml per well, and allowed to solidify for 30 minutes at room temperature. After this second overlay, plates were incubated at 37°C in 5% CO_2_ and observed for plaque formation, with final plaques counted on day three post second overlay. PRNT_90_ titres were defined as the inversed of the serum dilution at which plaque formation was reduced by 90%.

### Statistical analysis

Patients were categorised by district of residence, sex (female, male) and age group (<15 years, 15 – 54 years, and ≥ 55 years). Categorical variables were summarised using frequencies and percentages, while continuous variables, such as time from symptom onset to presentation, were described using means and ranges. CHIKV IgM seroprevalence infections was calculated as the proportion of CHIKV IgM-positive cases among the total number of samples tested. To identify independent predictors of CHIKV / ONNV infection, univariable logistic regression analyses were first conducted. Variables with *P* value <0.2 were subsequently included in a multivariable logistic regression model. Final variable selection for the multivariable model was performed using a stepwise selection algorithm based on the Akaike Information Criterion. Temporal trends in annual CHIKV / ONNV prevalence and associations between infection status and symptom burden were also assessed. All analyses were conducted using R software (version 4.4.2).

## Results

### Sampling

A total of 2756 serum samples collected between 19^th^ November 2018 and 29^th^ December 2022, from febrile patients presenting to sentinel surveillance health facilities in Uganda were available for laboratory screening. The samples were distributed across 63 out of 146 districts in Uganda **Figure 2**. Of these,75% were from four districts namely Kasese (29%, n=793), Wakiso (22%, n=596), Buikwe (14%, n=373) and Masaka (10%, n=262).

### Prevalence of CHIKV and ONNV in Uganda

All samples tested negative for CHIKV by RT-qPCR assays. Two samples tested positive for ONNV. Anti-CHIKV IgM antibodies were found in 8.8% (n=235) of samples distributed across 34 of the 63 districts sampled **Supplementary Table 1.** Most anti-CHIKV IgM-positive cases were from Wakiso 52 (22%), Kasese 46 (20%), Masaka 33 (14%), Buikwe 30 (13%), and Bundibugyo 24 (10%) districts. The distribution of these cases varied across 128 sentinel health facilities. Majority of the cases were observed in Kisubi hospital 33 (14%), Bukakata health center 23 (11%), St. Francis Hospital, Nkokonjeru 22 (9%), Nyamirami health center 19 (8%) and Bundibugyo hospital 13 (6%). The six health facilities accounted for almost 50% of the anti-CHIKV IgM positive cases.

**Table 1:**
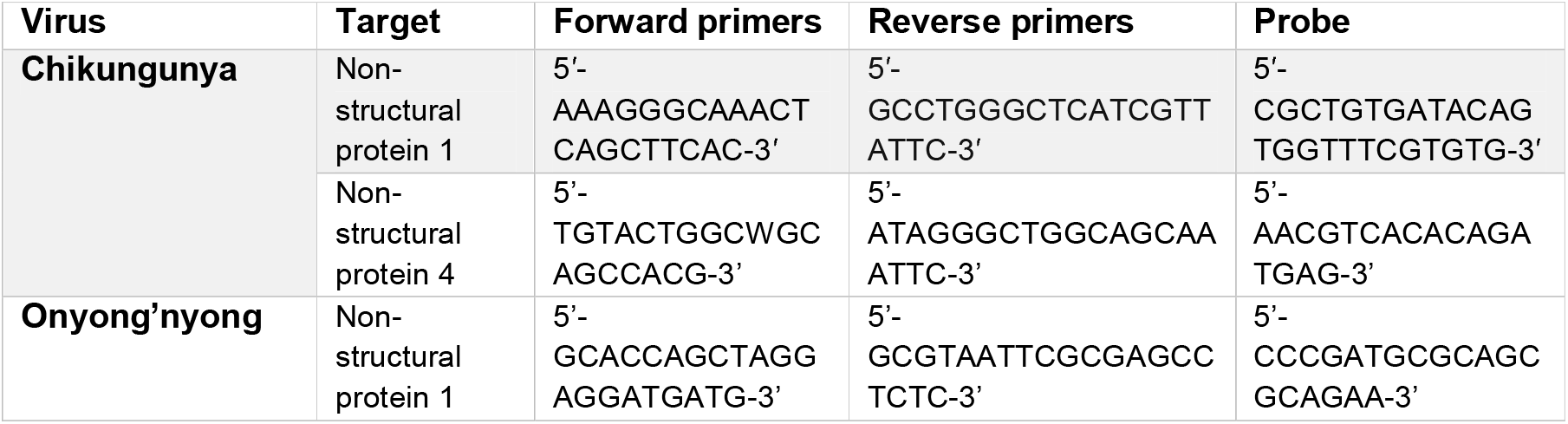
Primers and probes used for CHIKV and ONNV RT-PCR.

Anti-CHIKV IgM positivity occurred throughout the study period but with varying transmission intensities **Figure 3**. Most of the observed positive cases were reported in 2020 (20.9%), followed by 2021 (7.4%), then 2019 (5.9%) and 2021 (3.9%). There were no reported cases in 2018 and 2022 revealing a significant interannual variation in Anti-CHIKV IgM incidence (p < 0.001).

**Figure 3:**
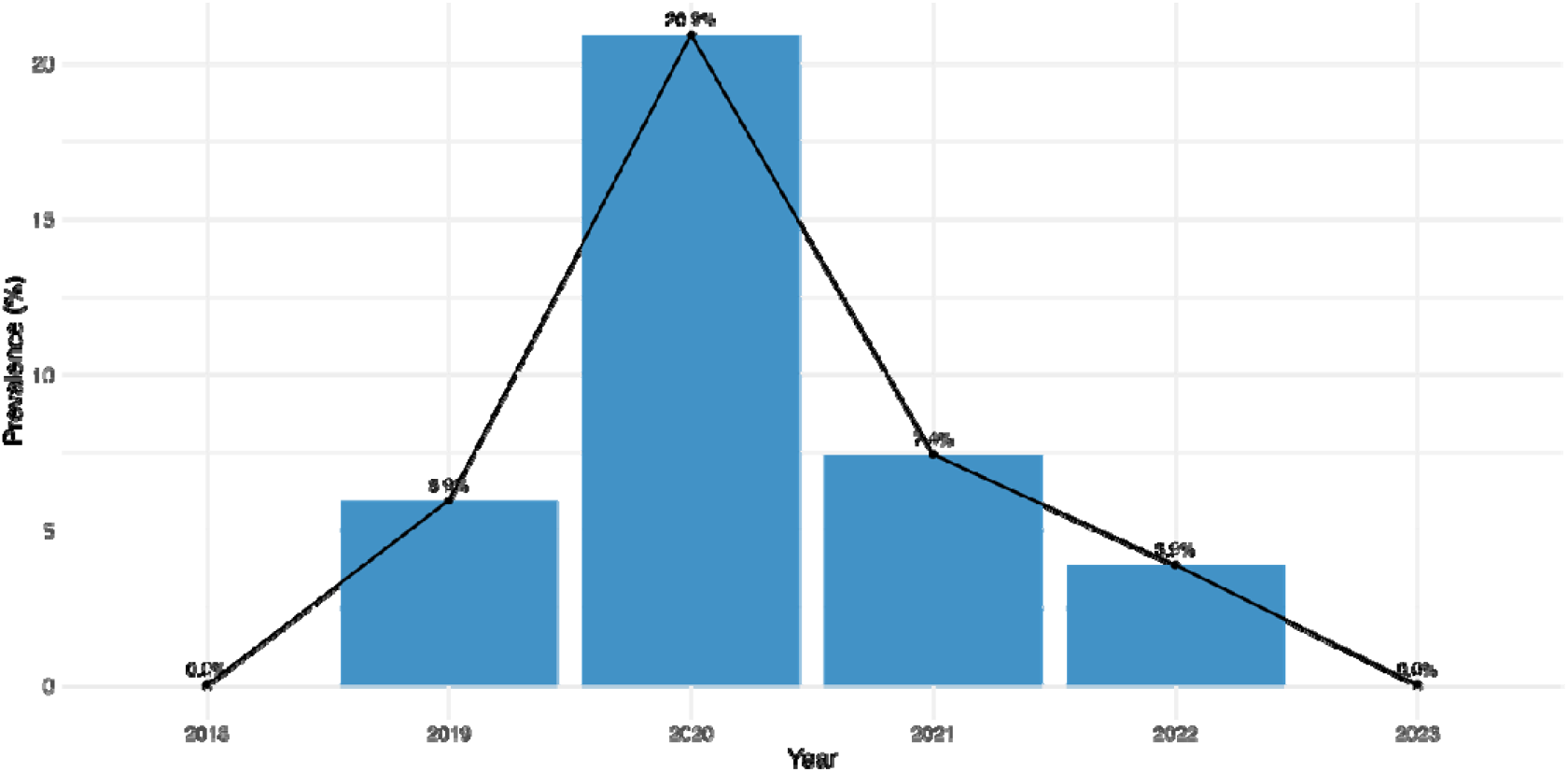
Prevalence of anti-CHIKV IgM positive cases over time in Uganda.

Anti-CHIKV IgM positivity variation across age groups was statistically significant (p = 0.02). Out of the 2756 patients, 11% (n = 316) were children below 15 years, 83% (n = 2281) were individuals of age 15 – 54 years and 6% (n = 159) were individuals of over 55 years **Table 2**. We observed the highest infection rate among children of age below 15 years (12.7%), followed by older adults of age 55+ with infection rate of 8.2% and 15-54 years (8.0%). **Table 2.**

**Table 2:** Distribution of epidemiological and clinical features of CHIKV/ONNV infections in Uganda.

|  | Characteristics | Negative(N=1183) | Positive(N =158) | Total(N=1341) | p.value |
| --- | --- | --- | --- | --- | --- |
| 1 | Gender |  |  |  | 0.819 |
| 2 | Male | 543 (45.9%) | 71 (44.9%) | 614 (45.8%) |  |
| 3 | Female | 640 (54.1%) | 87 (55.1%) | 727 (54.2%) |  |
| 4 | Year |  |  |  | < 0.001 |
| 5 | 2019 | 334 (28.2%) | 26 (16.5%) | 360 (26.8%) |  |
| 6 | 2020 | 326 (27.6%) | 92 (58.2%) | 418 (31.2%) |  |
| 7 | 2021 | 447 (37.8%) | 35 (22.2%) | 482 (35.9%) |  |
| 8 | 2022 | 76 (6.4%) | 5 (3.2%) | 81 (6.0%) |  |
| 9 | Age category |  |  |  | 0.237 |
| 11 | <15 | 253 (21.5%) | 42 (26.6%) | 295 (22.1%) |  |
| 12 | 15-54 | 807 (68.7%) | 105 (66.5%) | 912 (68.4%) |  |
| 13 | 55 | 115 (9.8%) | 11 (7.0%) | 126 (9.5%) |  |
| 14 | Symptoms |  |  |  |  |
| 15 | Fever | 883 (74.6%) | 132 (83.5%) | 1015 (75.7%) | 0.014 |
| 16 | Abdominal pain | 731 (61.8%) | 110 (69.6%) | 841 (62.7%) | 0.056 |
| 17 | Muscle pain | 661 (55.9%) | 89 (56.3%) | 750 (55.9%) | 0.914 |
| 18 | Skin Rash | 129 (10.9%) | 16 (10.1%) | 145 (10.8%) | 0.767 |
| 19 | Joint Pain | 730 (61.7%) | 92 (58.2%) | 822 (61.3%) | 0.399 |
| 20 | Headache | 848 (71.7%) | 101 (63.9%) | 949 (70.8%) | 0.044 |
| 21 | Vomit/Nausea | 597 (50.5%) | 80 (50.6%) | 677 (50.5%) | 0.976 |
| 26 | Temperature |  |  |  | 0.133 |
| 27 | Mean (SD) | 37.9 (1.68) | 37.6 (2.93) | 37.9 (1.87) |  |
| 28 | Range | 1.0 - 43.0 | 2.0 - 40.0 | 1.0 - 43.0 |  |

### CHIKV and ONNV Cross-reactivity

We performed plaque reduction neutralisation tests (PRNT) on 185 out of 235 serum samples that had initially tested positive for anti-CHIKV IgM; an additional 50 samples were unavailable for PRNT due to prior depletion. Notably, 73% (135 of 185) of these samples did not show confirmatory neutralising activity, indicating a high rate of false positives. Among the 50 samples that demonstrated alphavirus neutralisation, 60% had higher neutralising antibody titres against O’nyong-nyong virus (ONNV) than against Chikungunya virus (CHIKV). This finding suggests substantial serological cross-reactivity and the potential misclassification of ONNV infections as CHIKV. The remaining 20 samples demonstrated evidence of alphaviral neutralisation but there was no significant difference between the neutralisation antibody titres for CHIKV and ONNV **Figure 4**.

**Figure 4:**
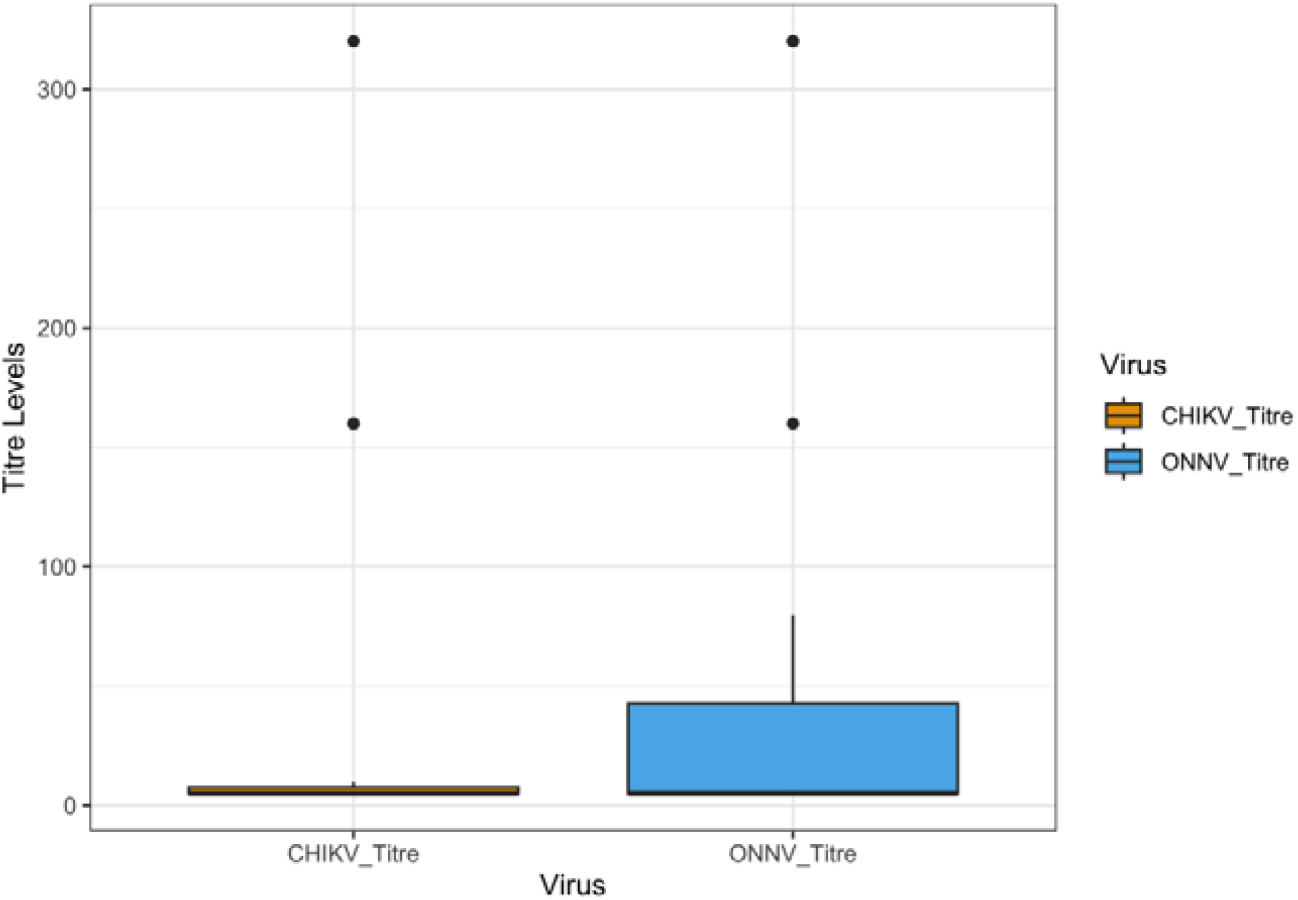
Distribution of CHIKV and ONNV neutralising titers among CHIKV IgM positive samples.

### Clinical characteristics associated with CHIKV IgM positive cases

Among patients who tested positive for anti-CHIKV IgM, most had fever (58%), abdominal pain (49%), loss of appetite (47%), headache (46%), general weakness (46%), joint pain (42%), muscle pain (40%). However, a significant proportion had fever (84.0%) compared to those who were negative (75.2%) with (p = 0.012). Other common symptoms among the positive cases were headache (65.3%), joint pain (59.3%), muscle pain (56.4%) compared to the negative groups, although all were not significantly different. There were no significant differences in skin rash, eye pain and bleeding among the positive and negative cases **Figure 5, Table 2.** Infected cases had a significant higher average number of symptoms with a mean of 2.1 as compared to negative cases with a mean of 1.6 (p < 0.001). Admission status was reported in 337 of the 2756 patients. The mean duration of symptom onset to admission did not differ significantly between positive and negative cases (median 4.2 vs 4.4 days, p = 0.825).

**Figure 5:**
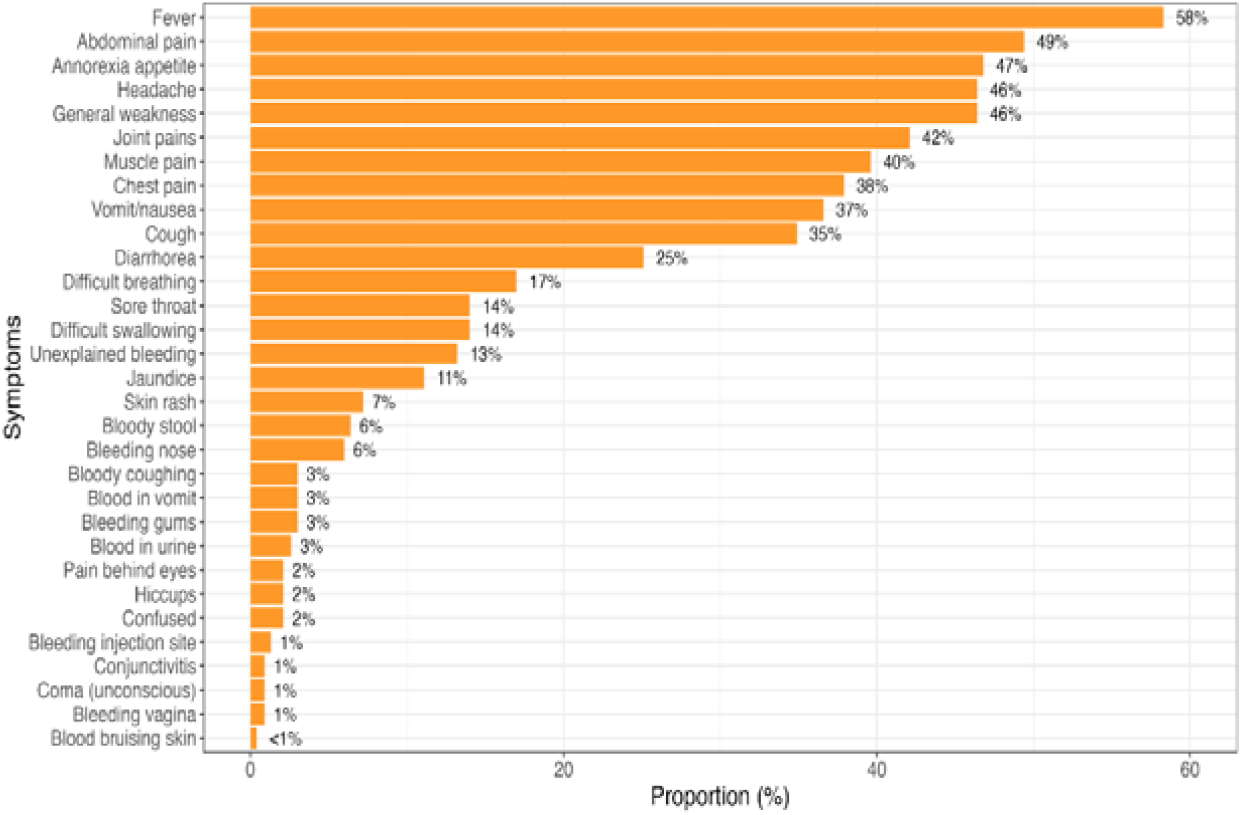
Distribution of symptoms among anti-CHIKV IgM positive cases.

Compared to anti-CHIKV IgM negative cases, a higher proportion of patient admissions occurred among CHIKV IgM positive cases (37 cases, 23%). Infected patients aged 15 years and below were admitted less frequently as compared to negative cases, suggesting that admission to hospitals was not attributable to anti-CHIKV IgM positivity. However, those aged 15 years and above were admitted more frequently compared to negative patients suggesting that admissions were attributable to anti-CHIKV IgM positivity.

There was also no significant difference based on the seasonality between the two groups; however, most of the positive cases were observed in dry season 179 (76.2%) compared to wet season 56 (23.8%).

### Risk factor analysis

The variables significantly associated with anti-CHIKV IgM positivity included Fever, year, abdominal pain, temperature and headache. A logistic regression model was applied to determine the relationship between the selected variables and infection **Figure 6.**

**Figure 6:**
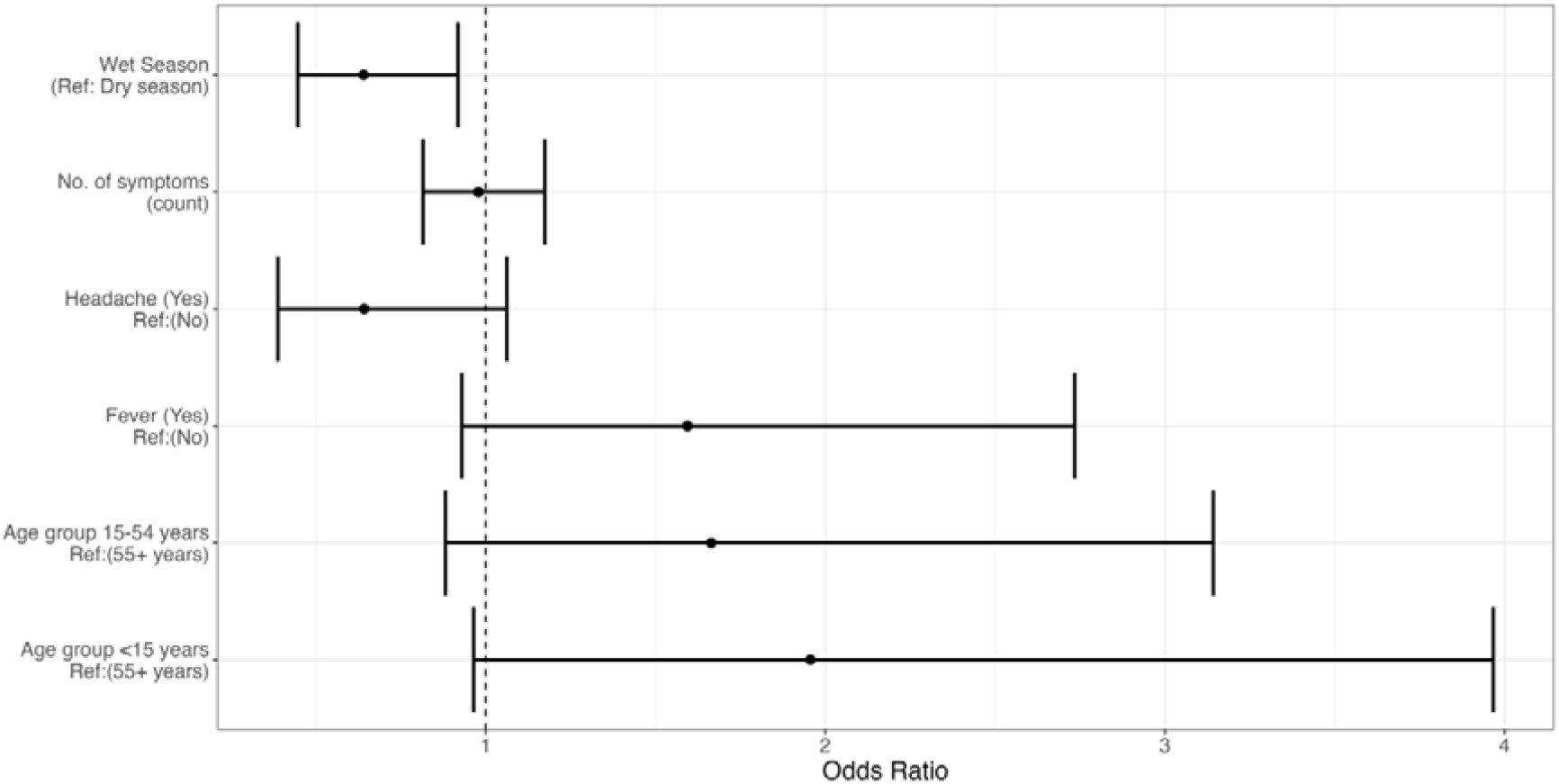
Factors associated with alphaviral infection in Uganda.

The results show that having fever increases the chances of anti-CHIKV IgM positivity though the relationship is not statistically significant. There is a stronger association between the year and positivity. Compared to the reference year (2019), 2020 had a higher odd of 3.86. This suggests a higher circulation of CHIKV/ONNV in 2020 compared to the baseline year. However, there was no significant difference between the baseline year and 2021 as well as 2022.

There was a positive association between abdominal pain and anti-CHIKV IgM positivity though the relationship was not statistically significant. Presence of abdominal pain increases the chances of anti-CHIKV IgM positivity by 45%. There was negative association between headache and anti-CHIKV IgM positivity. This means that the symptom may reflect a non-specific symptom or may be more prominent in other febrile illness than in CHIKV/ONNV infection. The association between temperature and anti-CHIKV IgM positivity was not statistically significant.

We performed a stepwise logistic regression model to select the most relevant variables to anti-CHIKV IgM infection. The stepwise logistic regression model identified fever as the strongest predictor of CHIKV/ONNV infection **Figure 7**.

**Figure 7:**
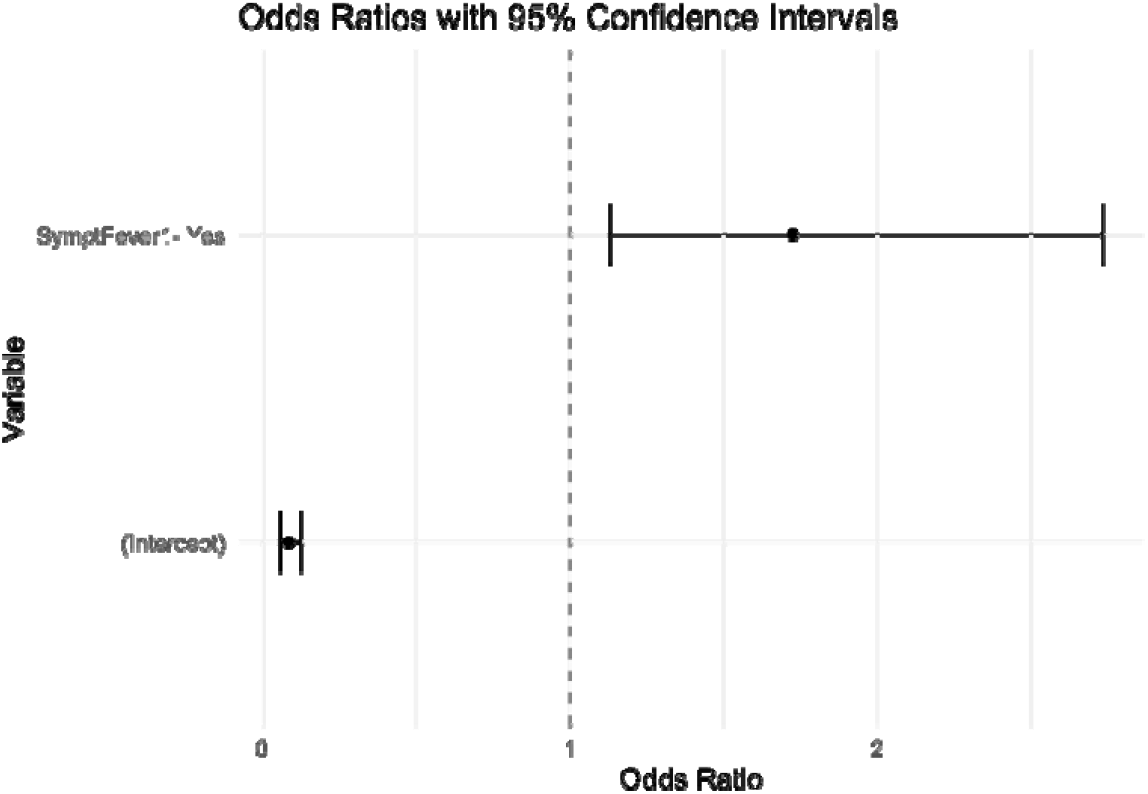
Individuals who reported fever had significantly higher odds of testing positive for anti-CHIKV IgM compared to those without fever.

## Discussion

This study presents the most comprehensive assessment to date of CHIKV and ONNV infections in Uganda, based on samples collected through nationwide sentinel surveillance over a five-year period (2018–2022). The findings offer new insight into the ongoing transmission and under-recognised burden of CHIKV and ONNV infections in Uganda. Notably, we found no molecular evidence of CHIKV in all the 2,756 samples tested. However, we confirmed ONNV infection in two individuals, each representing different occupations and geographic regions, suggesting broader, silent circulation. These results align with historical accounts identifying Uganda as the epicentre of ONNV outbreaks^15,24^ and further underscore the virus’s continued but largely undetected presence. Notably, our findings are also consistent with recent data from Rwanda, where farmers were found to be at higher risk of ONNV infection compared to those in other occupations ^25^.

Although low PCR positivity is common in arboviral surveillance due to the short duration of viraemia, our findings may also reflect sporadic ONNV circulation or missed detection linked to the timing of sample collection. In Uganda, health-seeking behaviours often result in delayed presentation to healthcare facilities, meaning many patients are likely sampled outside the narrow window of detectable viraemia. This poses a particular challenge for identifying infections caused by viruses like CHIKV and ONNV, which are characterised by brief periods of viraemia. Similar findings have been reported in Burkina Faso, where no alphaviruses were detected by RT-PCR despite serological evidence of recent infections ^26^, further underscoring the limitations of relying solely on molecular diagnostics in these settings.

Serological screening showed that 8.8% of participants had IgM antibodies against CHIKV, suggesting recent infection. However, PRNT revealed that most of these CHIKV IgM-positive samples had stronger neutralising responses to ONNV than to CHIKV. This indicates substantial serological cross-reactivity and raises the likelihood of misclassification when relying solely on IgM-based diagnostics. These findings, consistent with previous reports ^20,26,27^, highlight the importance of incorporating confirmatory neutralisation assays in endemic regions where multiple alphaviruses co-circulate, to ensure accurate diagnosis and disease burden estimation.

Temporal analysis in this study revealed a notable spike in anti-CHIKV IgM positivity in 2020, suggesting a previously unrecognised surge in alphavirus transmission during that period. This trend appears to align with known seasonal patterns of arboviral activity, which are often driven by changes in vector abundance and environmental conditions particularly during the dry season ^12,28^. Supporting this, surveillance data from Rwanda also reported ONNV IgM positivity in 2020, specifically among patients presenting to health facilities during the long dry season ^25^, further reinforcing the potential link between seasonal ecology and ONNV transmission dynamics.

The highest infection rates were recorded among children under 15 years, indicating early exposure and heightened susceptibility likely due to limited prior immunity. This pattern aligns with previous findings from coastal Kenya, where CHIKV disproportionately affected children ^21,22^ and from Rwanda, where similar trends were observed with ONNV ^25^. These observations highlight the need to prioritise this age group in the development and deployment of vaccines and other therapeutic interventions for effective control of alphaviral infections.

Clinically, fever was the most common and significant predictor of alphavirus infection, consistent with prior reports ^16,29^. Although symptoms like abdominal pain, headache, and joint pain were also common, they were not statistically distinct between anti-CHIKV IgM - positive and negative groups, highlighting the nonspecific clinical presentation of alphaviral illnesses and the challenge in differential diagnosis in resource-limited settings. Interestingly, we observed a higher admission rate among adults aged _≥_15 years who tested positive for anti-CHIKV IgM, suggesting greater disease severity in this age group.

This study is among the few to report real-world data on both CHIKV and ONNV in Uganda and provides evidence of the silent but continued circulation of ONNV. It highlights the limitations of current diagnostic tools and the critical need to develop more specific, multiplex platforms that can distinguish between co-circulating alphaviruses. Moreover, given the potential for ONNV to re-emerge in large outbreaks, as seen in the past, our findings call for renewed attention to this historically neglected virus.

### Study Limitations

This study had several limitations. First, the use of archived serum samples may have affected RNA integrity due to repeated freeze-thaw cycles, potentially impacting the sensitivity of molecular assays. Second, molecular detection was constrained by the timing of sample collection, many patients likely presented outside the narrow window of viraemia, reducing the likelihood of PCR positivity. Third, the absence of an ONNV-specific IgM ELISA may have affected the accuracy of seroprevalence estimates, as initial screening was conducted using CHIKV-based serological tools, which are known to cross-react with ONNV. Future studies should incorporate prospective sampling, alongside entomological and clinical data, to better characterise ONNV transmission dynamics and exposure risk. This integrative approach will be critical for informing the development of accurate diagnostics, targeted vector control, and potential vaccine or therapeutic strategies.

## Conclusion

This study reveals a significant and underrecognized burden of ONNV in Uganda, challenging long-held assumptions about the predominance of CHIKV in the region. The findings underscore the critical need to strengthen diagnostic capacity, particularly through the integration of confirmatory testing such as neutralisation assays into routine serological surveillance. The observed serological cross-reactivity highlights the risk of misclassification and the limitations of relying solely on IgM-based diagnostics.

Given ONNV’s resurgence potential, clinical overlap with CHIKV, and likely endemic transmission in vulnerable rural populations, there is an urgent need to prioritise ONNV within broader arboviral epidemic preparedness efforts. This includes investment in more sensitive, specific, and affordable diagnostic tools, as well as the incorporation of ONNV into vaccine development pipelines and vector control strategies. Ultimately, recognising ONNV’s role in febrile illness and integrating it into public health planning will be essential for reducing disease burden and improving epidemic response across East Africa.

### Institutional Review Board Statement

As the data were collected from patients as part of routine health care delivery and surveillance system for early warning of diseases and were anonymous, the Uganda Ministry of Health guidelines did not require approval by the UVRI Safety and Ethics Committee (SEC).

### Informed Consent Statement

Informed (oral) consent was obtained from all participants and their parents or legal guardians for those under the age of 16 years. All data was fully anonymized.

## Data Availability

All data produced in the present work are contained in the manuscript

