## Supplementary Table 1 for "The Burden of Chikungunya and Onyong’nyong Viral Infections in Uganda: A Hospital-Based Sentinel Surveillance Study"

**Supplementary table 1: Distribution of anti-CHIKV IgM positive cases per district**

| **District** | **Total cases** | **positive cases** | **prevalence(%)** |
| --- | --- | --- | --- |
| Wakiso | 397 | 46 | 11.6 |
| Kasese | 380 | 23 | 6.1 |
| Buikwe | 175 | 21 | 12.0 |
| Masaka | 145 | 25 | 17.2 |
| Bundibugyo | 111 | 17 | 15.3 |
| DRC | 35 | 2 | 5.7 |
| Mukono | 31 | 4 | 12.9 |
| Kagadi | 26 | 4 | 15.4 |
| Moyo | 21 | 0 | 0.0 |
| Buliisa | 16 | 4 | 25.0 |
| Arua | 13 | 2 | 15.4 |
| Kampala | 12 | 2 | 16.7 |
| Bunyangabo | 12 | 1 | 8.3 |
| Kikuube | 10 | 1 | 10.0 |
| Maracha | 10 | 0 | 0.0 |
| Kyegegwa | 9 | 2 | 22.2 |
| Kabale | 8 | 0 | 0.0 |
| Ntoroko | 8 | 0 | 0.0 |
| Kanungu | 7 | 2 | 28.6 |
| Yumbe | 7 | 0 | 0.0 |
| Kalangala | 6 | 2 | 33.3 |
| Isingiro | 6 | 0 | 0.0 |
| Adjumani | 5 | 1 | 20.0 |
| Kamwenge | 5 | 1 | 20.0 |
| Nebbi | 5 | 1 | 20.0 |
| Kakumiro | 4 | 1 | 25.0 |
| Kiboga | 4 | 1 | 25.0 |
| Mubende | 4 | 0 | 0.0 |
| Zombo | 3 | 1 | 33.3 |
| Kisoro | 3 | 0 | 0.0 |
| Kyankwanzi | 3 | 0 | 0.0 |
| Lyantonde | 3 | 0 | 0.0 |
| Mbarara | 3 | 0 | 0.0 |
| Nakaseke | 3 | 0 | 0.0 |
| Obongi | 3 | 0 | 0.0 |
| Rubanda | 3 | 0 | 0.0 |
| Tororo | 3 | 0 | 0.0 |
| Gulu | 2 | 0 | 0.0 |
| Kabarole | 2 | 0 | 0.0 |
| Kalungu | 2 | 0 | 0.0 |
| Kibaale | 2 | 0 | 0.0 |
| Kyotera | 2 | 0 | 0.0 |
| Lira | 2 | 0 | 0.0 |
| Masindi | 2 | 0 | 0.0 |
| Buvuma | 1 | 1 | 100.0 |
| Hoima | 1 | 1 | 100.0 |
| Kiryandongo | 1 | 1 | 100.0 |
| Mityana | 1 | 1 | 100.0 |
| Bukomansimbi | 1 | 0 | 0.0 |
| Bushenyi | 1 | 0 | 0.0 |
| Busia | 1 | 0 | 0.0 |
| Butebo | 1 | 0 | 0.0 |
| Kaliro | 1 | 0 | 0.0 |
| Kyenjojo | 1 | 0 | 0.0 |
| Luwero | 1 | 0 | 0.0 |
| Mayuge | 1 | 0 | 0.0 |
| Mpigi | 1 | 0 | 0.0 |
| Nakasongola | 1 | 0 | 0.0 |
| Pakwach | 1 | 0 | 0.0 |
| Rakai | 1 | 0 | 0.0 |
| Rukiga | 1 | 0 | 0.0 |
| Rwampara | 1 | 0 | 0.0 |
| Sembabule | 1 | 0 | 0.0 |
| Sheema | 1 | 0 | 0.0 |
